# Statistical Challenges when Analyzing SARS-CoV-2 RNA Measurements Below the Assay Limit of Quantification in COVID-19 Clinical Trials

**DOI:** 10.1101/2023.03.13.23287208

**Authors:** Carlee B Moser, Kara W Chew, Mark J Giganti, Jonathan Z Li, Evgenia Aga, Justin Ritz, Alexander L Greninger, Arzhang Cyrus Javan, Rachel Bender Ignacio, Eric S Daar, David A Wohl, Judith S Currier, Joseph J Eron, Davey M Smith, Michael D Hughes, ACTIV-2/A5401 Study Team

## Abstract

Most clinical trials evaluating COVID-19 therapeutics include assessments of antiviral activity. In recently completed outpatient trials, changes in nasal SARS-CoV-2 RNA levels from baseline were commonly assessed using analysis of covariance (ANCOVA) or mixed models for repeated measures (MMRM) with single-imputation for results below assay lower limits of quantification (LLoQ). Analyzing changes in viral RNA levels with singly-imputed values can lead to biased estimates of treatment effects. In this paper, using an illustrative example from the ACTIV-2 trial, we highlight potential pitfalls of imputation when using ANCOVA or MMRM methods, and illustrate how these methods can be used when considering values <LLoQ as censored measurements. Best practices when analyzing quantitative viral RNA data should include details about the assay and its LLoQ, completeness summaries of viral RNA data, and outcomes among participants with baseline viral RNA ≥LLoQ, as well as those with viral RNA <LLoQ.

**Trial Registration:** ClinicalTrials.gov Identifier: NCT04518410

## BACKGROUND

Clinical trials designed to evaluate COVID-19 therapeutics should have clinically meaningful endpoints. FDA guidance states that clinical outcomes, such as the proportion of participants hospitalized or time to symptom recovery, are recommended as primary outcomes in phase III outpatient COVID-19 trials [1]. However, it also states that viral shedding should be measured to assess antiviral activity, primary virology outcomes are acceptable in phase II, and quantitative and qualitative virological assessments are encouraged.

In typical COVID-19 randomized trials, samples such as nasopharyngeal swabs, anterior or mid-turbinate nasal swabs, oropharyngeal swabs, saliva, or plasma, are collected longitudinally for SARS-CoV-2 RNA testing before and after intervention. Repeat sampling from early timepoints is common and in phase III typically includes one to four timepoints (Supplemental Table 1).

To evaluate virologic efficacy, SARS-CoV-2 RNA, henceforth called viral RNA (vRNA), is measured with quantitative reverse transcription polymerase chain reaction (RT-qPCR) assays. Like other nucleic acid assays, SARS-CoV-2 RNA assays have limits between which vRNA is accurately quantified, called the lower limit of quantification (LLoQ) and upper limit of quantification (ULoQ). For results >ULoQ, samples can be rerun with dilution to obtain quantifiable values. Assays may also indicate whether results <LLoQ are detectable or not.

Recent outpatient COVID-19 therapeutic trials considered various vRNA outcome measures and statistical methods. Most commonly, vRNA changes from baseline were analyzed using analysis of covariance (ANCOVA) at each timepoint or mixed models for repeated measures (MMRM). With these methods, single-imputation was used to assign values for vRNA results <LLoQ (Supplemental Table 1) [2–18]. However, such imputation can introduce bias in estimating the magnitudes of treatment effects, as uncertainty for values <LLoQ isn’t captured [19].

Using an illustrative example from the ACTIV-2 COVID-19 outpatient treatment trial, we describe bias that may arise when estimating treatment effects using single-imputation with ANCOVA and MMRM. Drawing on the HIV literature [19], we describe and discuss alternative approaches for analyzing vRNA changes, that may be more appropriate by considering vRNA values <LLoQ as censored measurements. Finally, we provide recommendations for the analysis and presentation of results concerning vRNA changes in future trials.

## METHODS

ACTIV-2 (NCT04518410) is an adaptive platform trial designed to evaluate potential outpatient therapeutics for COVID-19[20]. Our illustrative example includes 114 participants randomized to receive tixagevimab/cilgavimab intravenously or placebo; the primary results previously reported [21]. Nasopharyngeal swabs were collected before treatment at Day 0 (baseline) and Days 3, 7 and 14 for SARS-CoV-2 RNA quantitative testing using a RT-qPCR assay with LLoQ of 2 log_10_ copies/ml [22]. All results >ULoQ were rerun with dilution to obtain quantifiable results. ACTIV-2 was approved by a central institutional review board (IRB), Advarra (Pro00045266), with additional local IRB review and approval as required by participating sites. All participants provided written informed consent.

As this manuscript aims to illustrate and discuss different approaches to analyze vRNA changes, we provide an overview in Table 1, but integrate descriptions of each method in the Results. For methods that use imputed values for results <LLoQ, two commonly-used single-imputation strategies (Supplemental Table 1) were assessed:

**Table 1:**
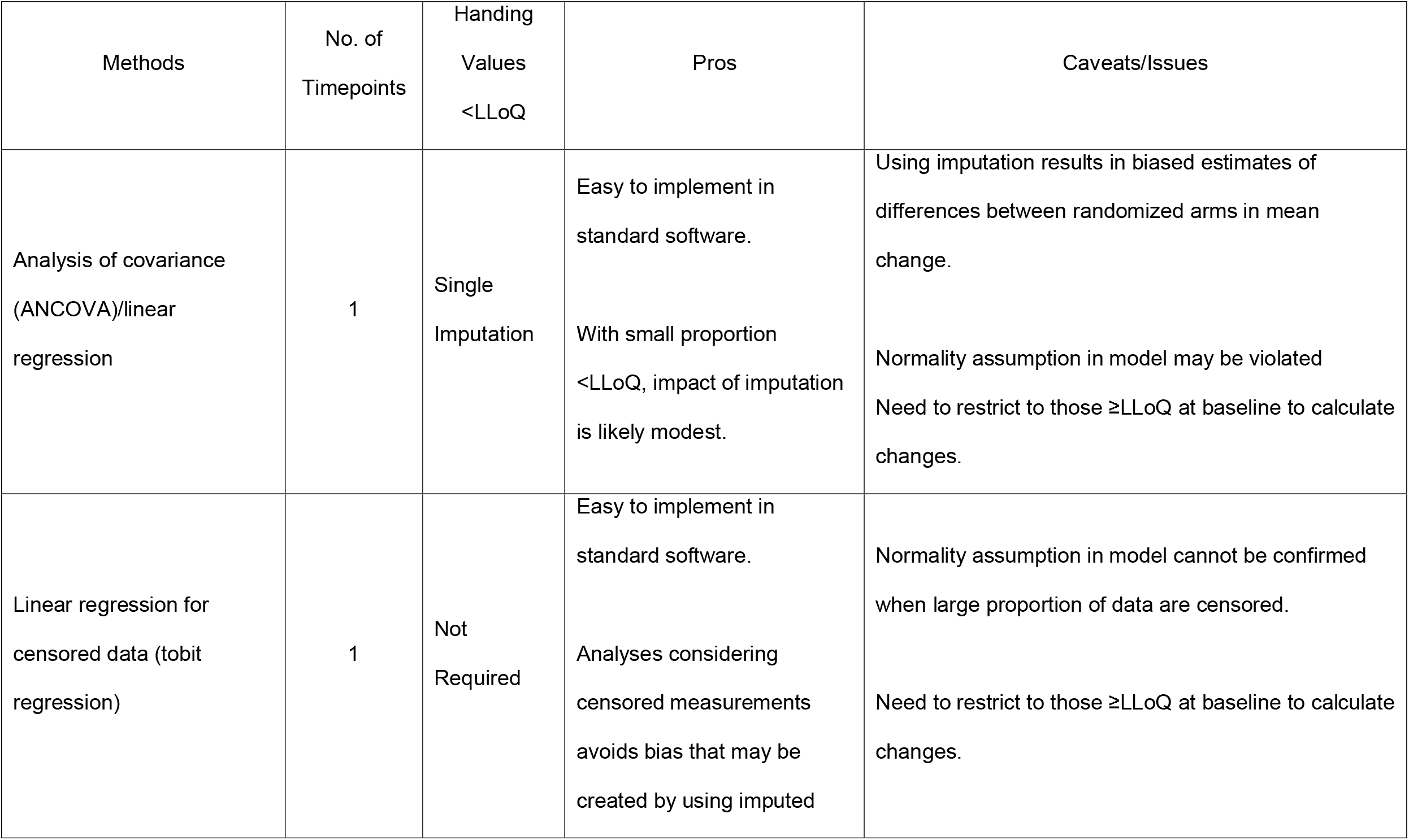
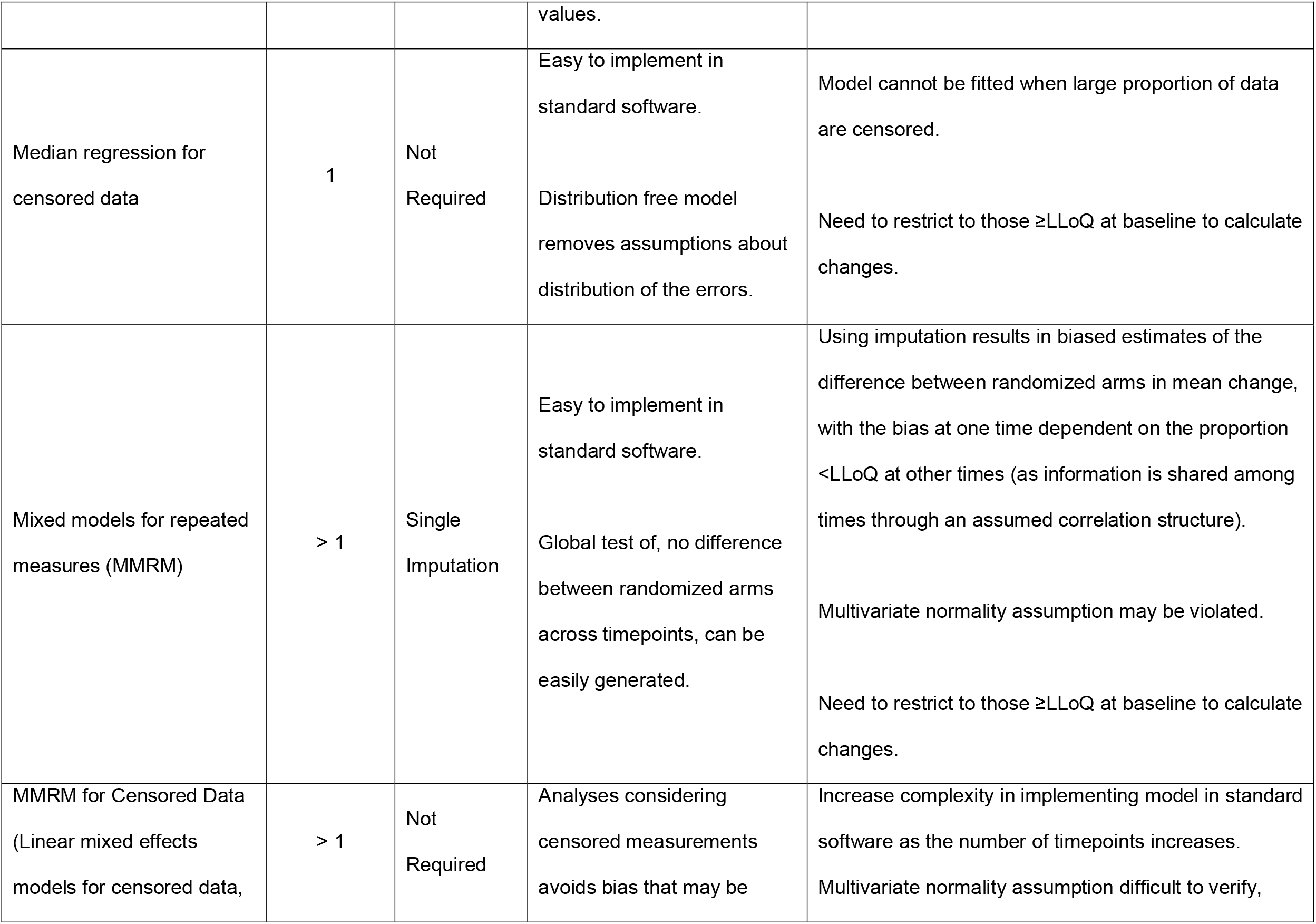
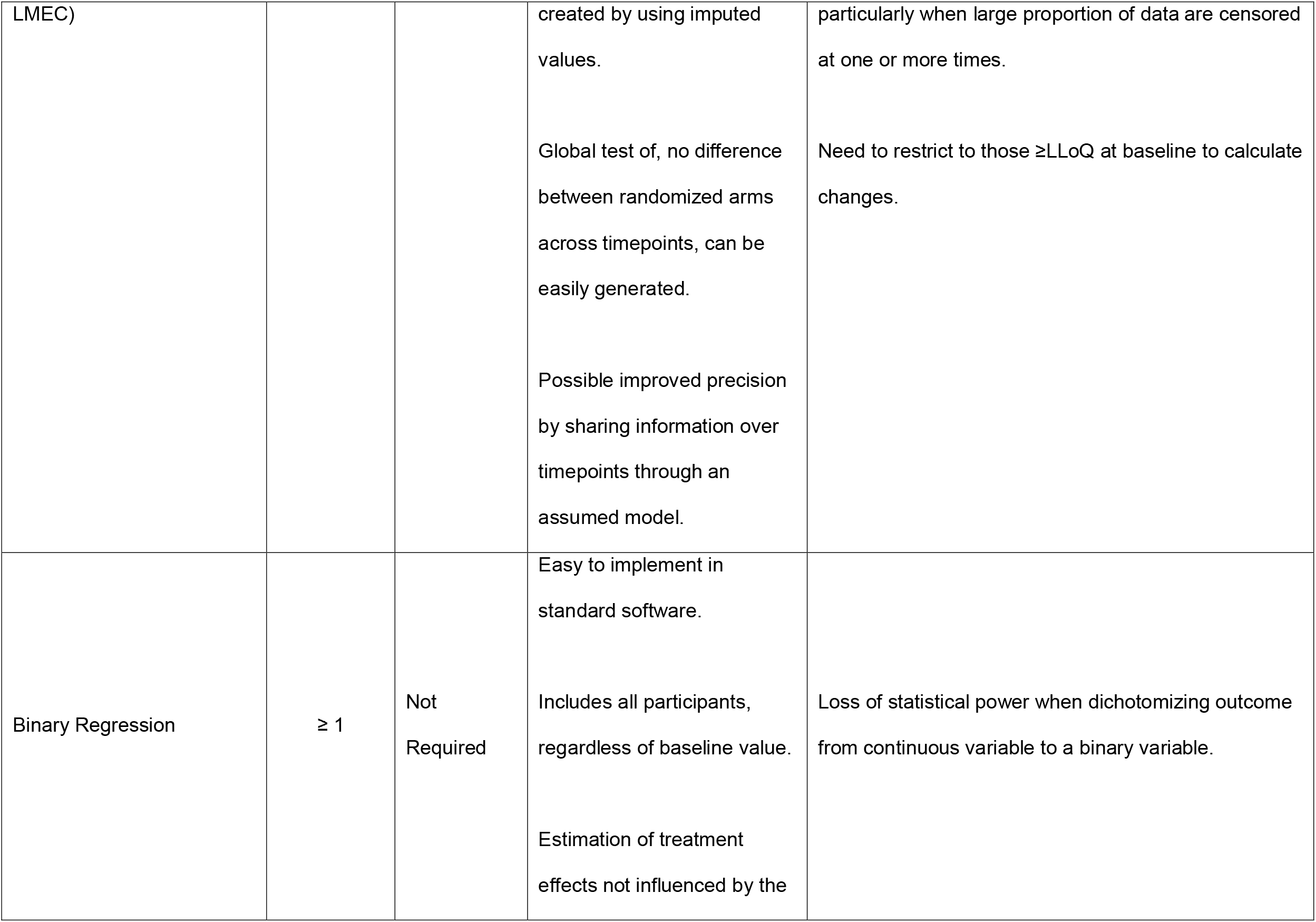
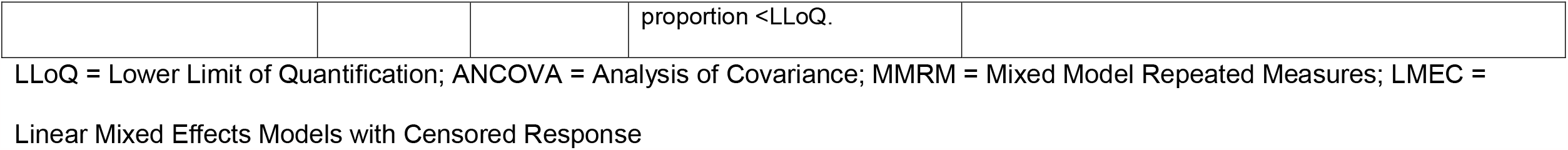
Summary of Analytic Methods Considered in Our Illustrative Example for the Analysis of Changes from Baseline in SARS-CoV-2 RNA proportion <LLoQ.

(1) “*LLoQ-imputation*”: impute values <LLoQ as the LLoQ,
(2) “*½LLoQ-imputation*”: impute values <LLoQ as ½ the LLoQ.

See Supplemental Methods for additional details on model specifications and sample SAS software code.

## RESULTS

Descriptive summaries of vRNA across timepoints for the 114 participants are shown in Table 2A and Figure 1A and 1B. At baseline, 15 participants (13%) had missing vRNA (Supplemental Figure 1). There was a chance imbalance in vRNA between the randomized arms, with median vRNA in the active arm 1.0 log_10_ copies/ml higher than the placebo arm, and a higher proportion of participants with vRNA ≥LLoQ (72% versus 62%).

**Table 2:** Distribution of SARS-CoV-2 RNA by Study Visit in each Treatment Arm in overall cohort (A) and among those with vRNA ≥LLoQ at Baseline/Day 0 (B)

| A: All participants in cohort (N=114) |  |  |  |
| --- | --- | --- | --- |
| Visit |  | Active (N=58) | Placebo (N=56) |
| Baseline | Median (quartiles) | 4.0 (<LLoQ, 6.6) | 3.0 (<LLoQ, 5.9) |
|  | <LLoQ, n (%) | 14 (28) | 19 (38) |
|  | Missing, n | 9 | 6 |
| Day 3 | Median (quartiles) | <LLoQ (<LLoQ, 3.9) | <LLoQ (<LLoQ, 3.9) |
|  | <LLoQ, n (%) | 26 (52) | 24 (52) |
|  | Missing, n | 8 | 10 |
| Day 7 | Median (quartiles) | <LLoQ (<LLoQ, 2.2) | <LLoQ (<LLoQ, 2.2) |
|  | <LLoQ, n (%) | 37 (74) | 35 (71) |
|  | Missing, n | 8 | 7 |
| Day 14 | Median (quartiles) | <LLoQ (<LLoQ, <LLoQ) | <LLoQ (<LLoQ, <LLoQ) |
|  | <LLoQ, n (%) | 49 (98) | 45 (97) |
|  | Missing, n | 11 | 7 |
| B: All participants with vRNA ≥ LLoQ at baseline (N=66) | | | |
| Visit |  | Active (N=35) | Placebo (N=31) |
| Baseline | Median (quartiles) | 5.5 (3.7, 8.0) | 5.0 (3.1, 6.7) |
|  | <LLoQ, n (%) | 0 (0) | 0 (0) |
|  | Missing, n | 0 | 0 |
| Day 3 | Median (quartiles) | 3.0 (<LLoQ, 4.5) | 3.4 (<LLoQ, 5.9) |
|  | <LLoQ, n (%) | 8 (27) | 7 (28) |
|  | Missing, n | 5 | 6 |
| Day 7 | Median (quartiles) | <LLoQ (<LLoQ, 2.5) | <LLoQ (<LLoQ, 3.3) |
|  | <LLoQ, n (%) | 18 (62) | 14 (54) |
|  | Missing, n | 6 | 5 |
| Day 14 | Median (quartiles) | <LLoQ (<LLoQ, <LLoQ) | <LLoQ (<LLoQ, <LLoQ) |
|  | <LLoQ, n (%) | 27 (93) | 24 (89) |
|  | Missing, n | 6 | 4 |

**Figure 1:**
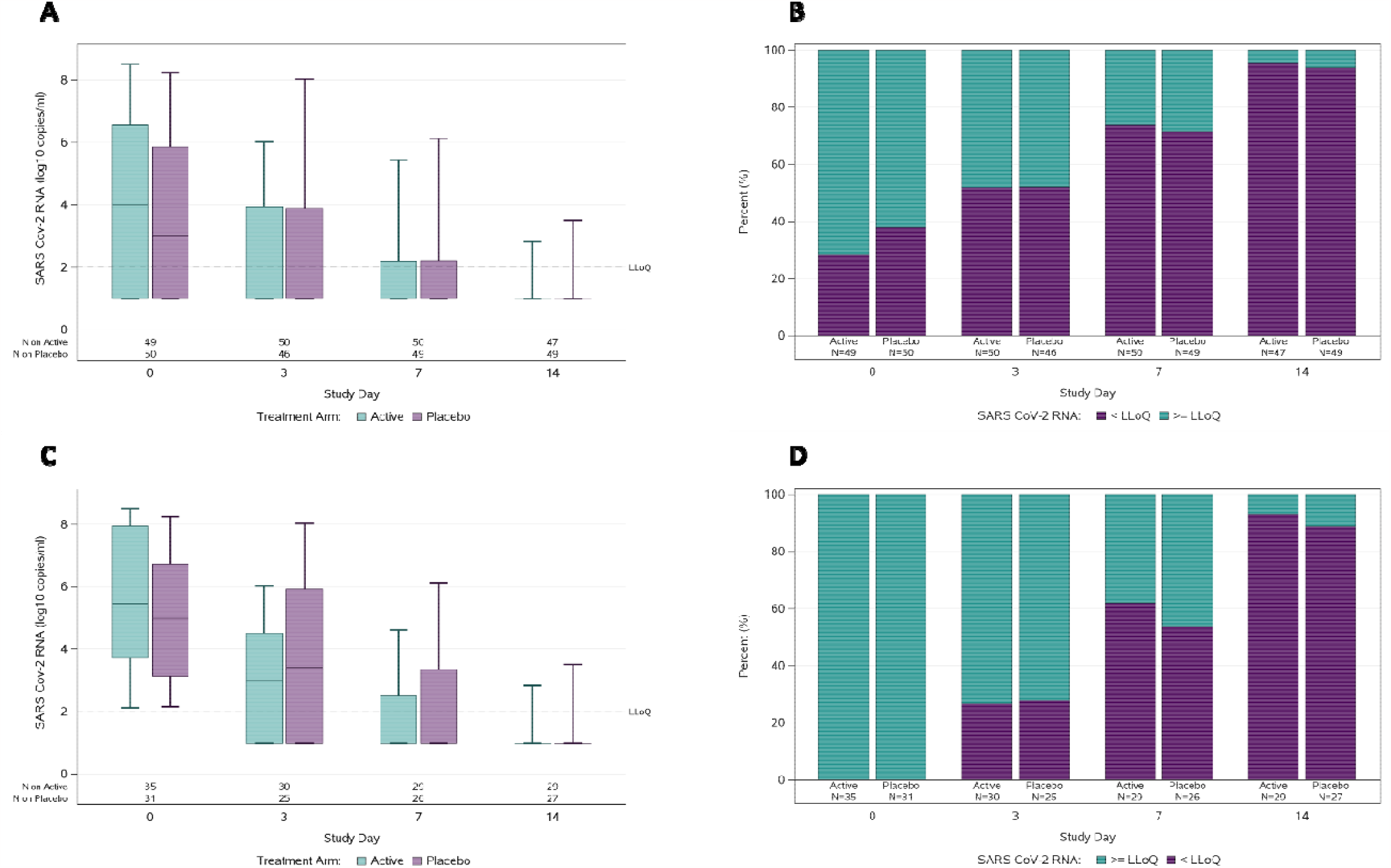
Distribution of SARS-CoV-2 RNA from nasopharyngeal swabs in Active and Placebo arms by study visit in overall cohort (A and B) and among those with vRNA ≥LLoQ at Baseline/Day 0 (C and D). Levels of SARS-CoV-2 RNA (log_10_ copies/ml) with horizontal line = median, box=interquartile range, whiskers = minimum/maximum (A and C); results below the LLoQ are plotted using an imputed value of 1 log_10_ copies/ml. Proportion with quantifiable SARS-CoV-2 RNA (green) and unquantifiable (purple) (B and D). LLOQ = Lower Limit of Quantification.

Following the recommendation of Marschner et al. [19], we separately considered data for participants with vRNA <LLoQ from those ≥LLoQ at baseline. For those with vRNA <LLoQ at baseline (N=33), vRNA remained <LLoQ at all follow-up timepoints in both arms, suggesting peak vRNA may have been achieved before enrollment. For the remaining analyses, we focus on the 66 participants with vRNA ≥LLoQ at baseline. The proportion with vRNA <LLoQ increased over time: 27% and 28% at Day 3, 62% and 54% at Day 7, and 93% and 89% at Day 14 for the active and placebo arms, respectively (Table 2B and Figure 1C and 1D).

### Analyzing vRNA at a Single Timepoint

#### 1. Using imputed values leads to biased estimates

Fifty-five (83%) of the 66 participants had vRNA results at Day 3 (Supplemental Figure 1). For these 55 participants, at baseline there was a modest difference (0.33 log_10_ copies/ml) in mean vRNA: 5.61 and 5.28 log_10_ copies/ml for the active and placebo arms, respectively.

Using *LLoQ-imputation*, the mean vRNA at Day 3 was 3.43 and 3.97 log_10_ copies/ml for the active and placebo arms, respectively, with estimated mean changes from baseline of -2.18 and -1.30 log_10_ copies/ml. Within each arm, the estimated mean changes are conservative and biased because for participants with vRNA <LLoQ at Day 3, the true changes are at least as large in magnitude as the imputed changes. Using *½LLoQ-imputation* gives mean changes that are larger (more negative) compared to *LLoQ-imputation*: -2.45 and -1.58 log_10_ copies/ml for the active and placebo arms, respectively. This imputation still results in biased estimates, but with an unknown direction (estimated changes may be larger or smaller than the truth). For both approaches, the larger mean change in the active arm could reflect higher average baseline values, and thus larger changes are observable. Since the estimated mean changes within each arm are biased, the estimated difference between arms will be biased, and further bias may be introduced with the baseline imbalances.

The estimated difference in mean change for the active versus placebo arms at Day 3 was -0.87 log_10_ copies/ml using *LLoQ-imputation* and -0.86 log_10_ copies/ml using *½LLoQ-imputation* (Table 3A). Although these estimates are similar, this may not be the case in other datasets when using the two approaches. By Day 14, when ∼90% of participants had vRNA <LLoQ (and hence had imputed changes), the estimated difference in mean change between arms was approximately equal to the baseline mean difference for both imputation approaches. If all participants had vRNA <LLoQ at Day 14, the difference in mean change would equal the difference in mean vRNA at baseline, despite the choice of imputed value and underlying true difference. With larger proportions <LLoQ, differences between arms can reflect chance imbalances at baseline rather than true differences.

**Table 3:**
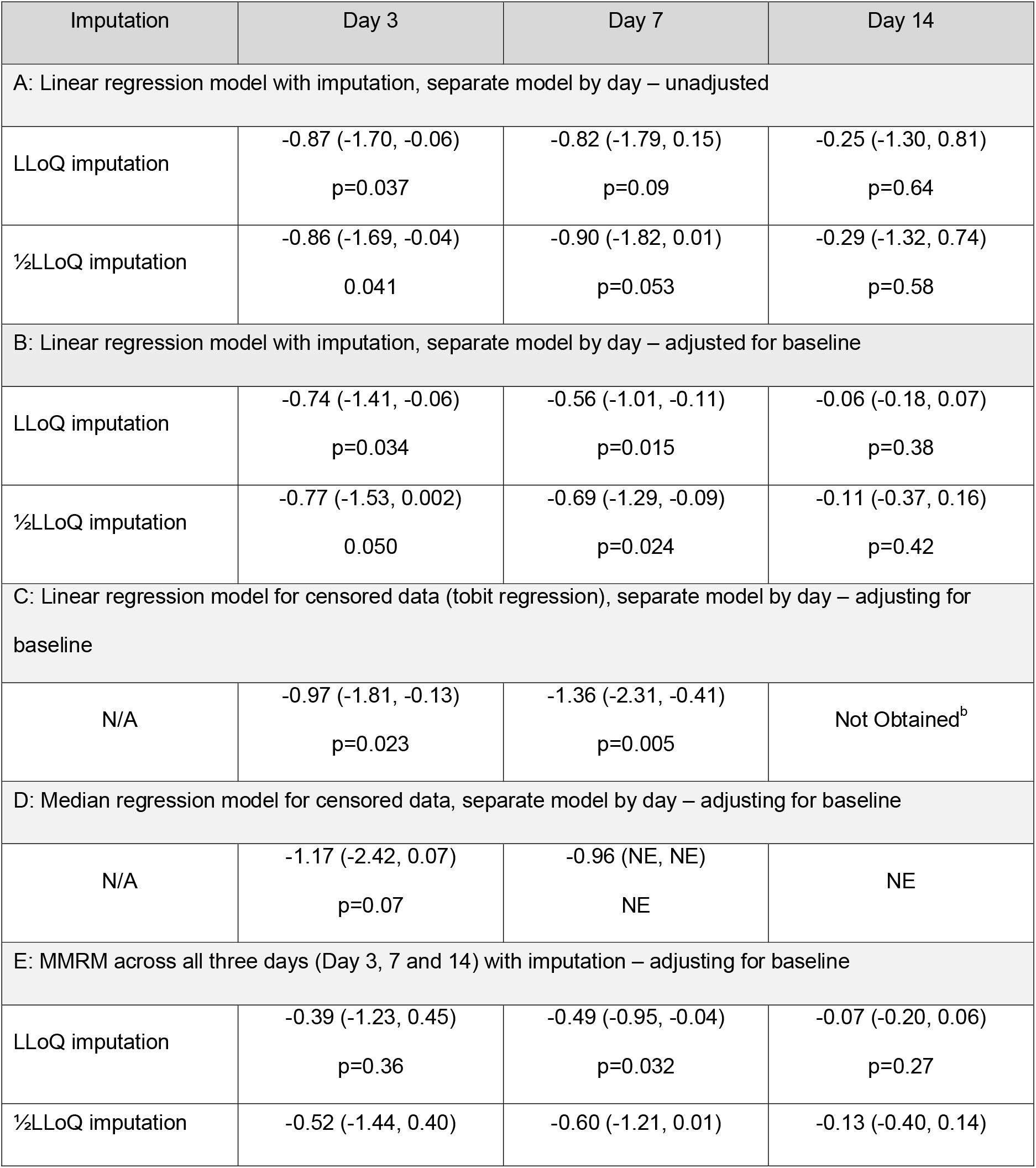
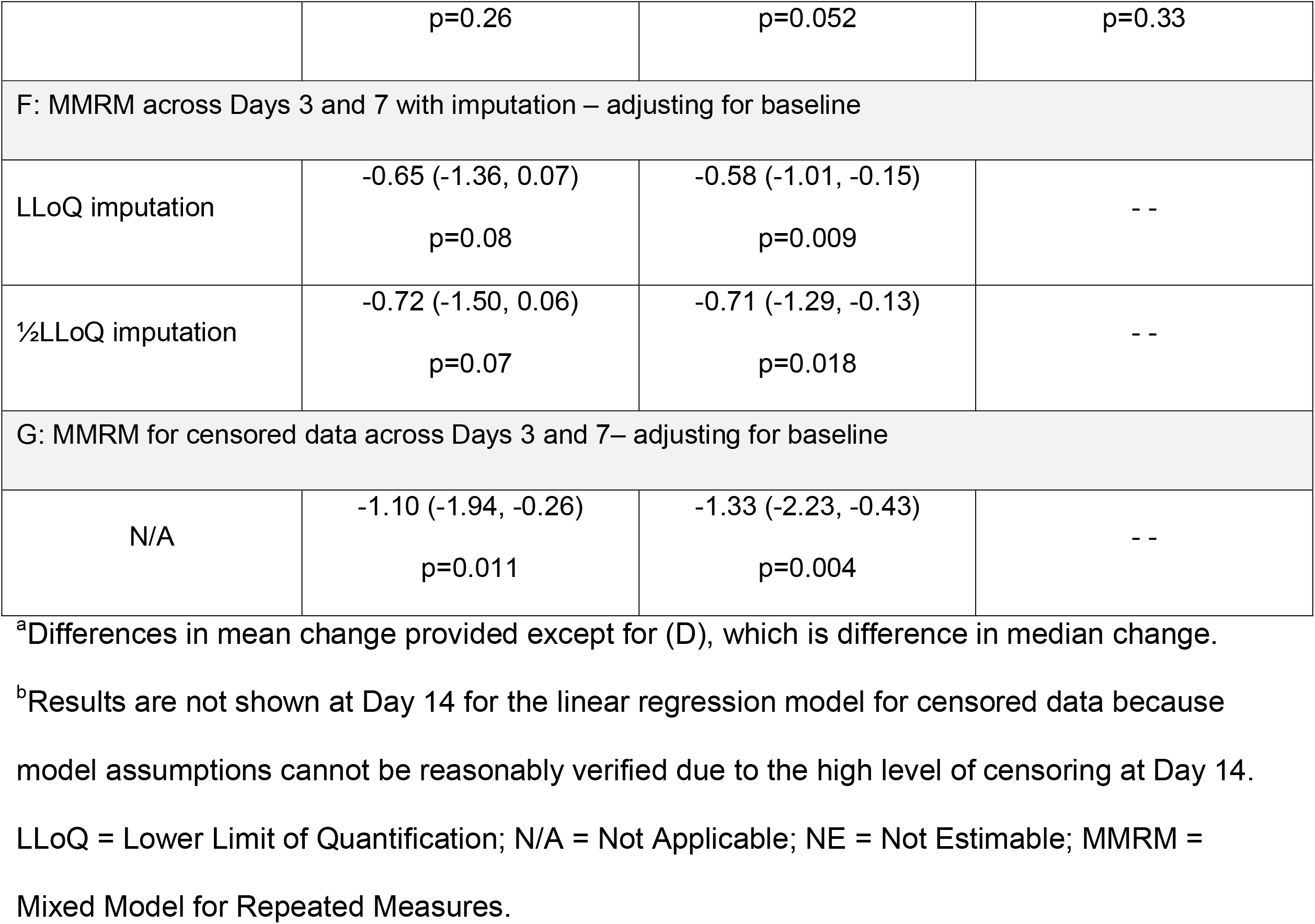
Differences between Treatment Arms in SARS-CoV-2 RNA (log_10_ copies/ml) change from baseline– Mean/Median^a^, 95% CI and p-value among those with quantifiable baseline vRNA

#### 2. Adjusting for baseline can help address baseline imbalances

Although adjusting for baseline doesn’t remove the bias in estimating differences between arms using singly-imputed values, it may help reduce the impact of baseline imbalances in mean vRNA when assessing treatment effects.

The estimated differences in mean changes between arms using standard linear regression are shown in Table 3A-B. In adjusted analyses, differences between arms have some attenuation at each timepoint compared with unadjusted analyses, reflecting the adjustment for higher baseline vRNA levels in the active arm.

#### 3. Analysis methods considering vRNA <LLoQ as censored

Statisticians refer to vRNA values <LLoQ as being left-censored because the if the true vRNA could be measure it would be a value between zero and LLoQ (i.e., a value to the left of LLoQ). This contrasts with right-censoring like in survival analysis where, for example, participants alive at the end of follow-up have time of death greater than (to the right of) the time at the end of follow-up. Statistical methods used for survival analysis can be used to analyze vRNA data, with the small adaptation that values are left-censored rather than right-censored. Change in vRNA is defined as the difference in vRNA at the follow-up time minus the baseline. However, for follow-up vRNA values that are <LLoQ or left-censored, the change in vRNA is calculated as the LLoQ minus baseline vRNA, and is also left-censored.

Linear regression using software designed to handle censored data (known as tobit regression) is a possible method. Using this approach, adjusting for baseline vRNA, the estimated difference between arms in mean change from baseline to Day 3 was -0.97 log_10_ copies/ml (95% confidence interval [CI]: -1.81, -0.13) favoring the active arm (Table 3C), and is somewhat larger than the differences in mean change by either imputation approach (Table 3B). At Day 7, the difference in mean change from baseline was -1.36 log_10_ copies/ml, also favoring the active arm (95% CI: -2.31, -0.41), which is much larger than differences observed by either imputation approach, illustrating the potential bias using those methods when the proportion with vRNA <LLoQ increases. We didn’t pursue an analysis of mean changes to Day 14 using tobit regression because of the high level of censoring (∼90%) and hence the inability to check model assumptions.

As with standard linear regression, there is an assumption that the errors in the model are normally distributed. These errors are estimated by the residuals calculated as the observed vRNA value minus the predicted model value. The distributional assumption can be evaluated with quantile-quantile (Q-Q) plots, comparing the quantiles of the observed distribution of the residuals (calculated using Kaplan-Meier methods to account for censored residuals) against the corresponding quantiles of a standard normal distribution. If the assumption was satisfied, the plots would show linear associations. Figure 2 shows Q-Q plots for the distribution of standardized residuals from models for change from baseline, adjusting for baseline. For the models of change from baseline to Days 3 and 7, the Q-Q plots appear reasonably linear, supporting normality assumptions. We note, however, the more restricted range of the Q-Q plot for changes to Day 7, as shown by the lack of standardized residuals below -1. This reflects the higher proportion of censored values at Day 7; thus, the normality assumption cannot be verified for the tail of the distribution, corresponding to large negative changes from baseline.

**Figure 2:**
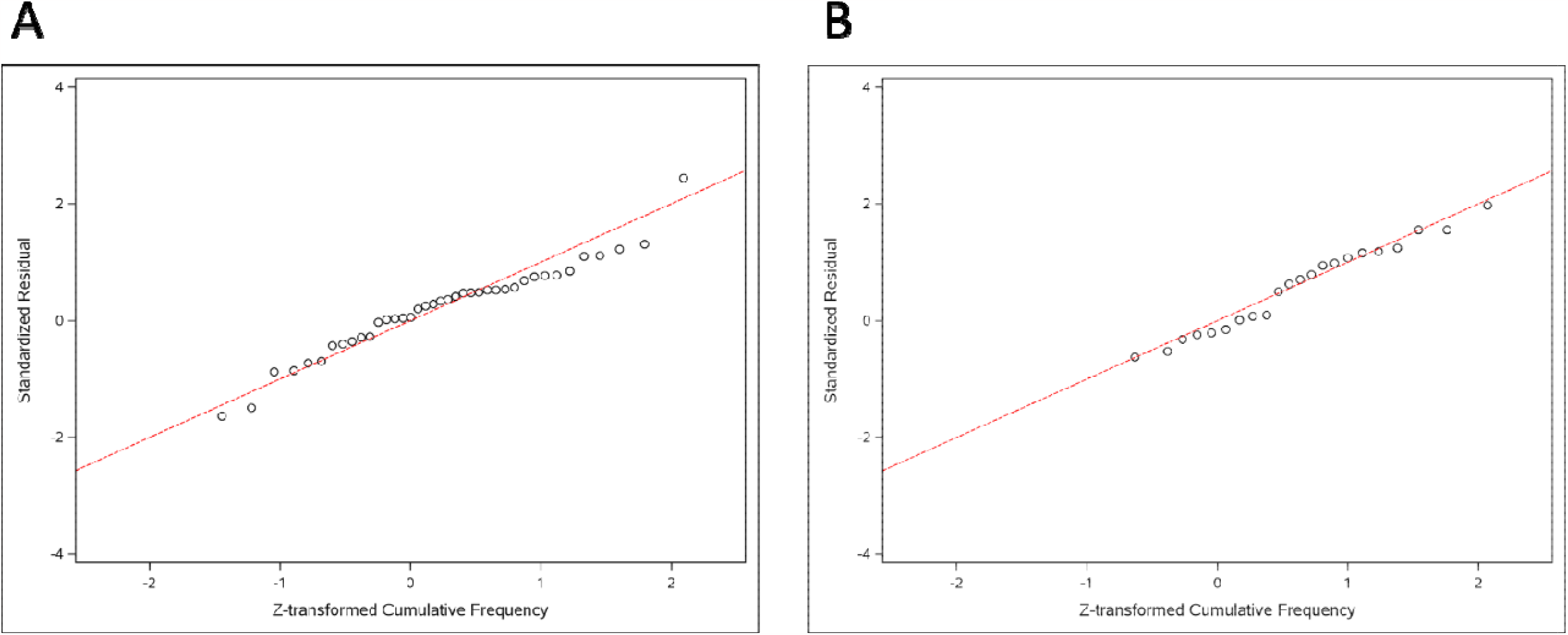
Quantile-quantile (Q-Q) plot for linear regression model for censored data for change in vRNA from baseline to Day 3 (A) and to Day 7 (B), both models included an indicator variable for treatment versus placebo and adjusted for baseline vRNA Standardized residuals (for the non-censored observations) calculated by dividing the residuals by their standard deviation (estimated from the fitted model). Quantiles for a standard normal distribution plotted on the x-axis take account of censored residuals.

#### 4. Quantile regression as an alternative distribution-free method

An alternative to tobit regression is quantile regression applied to assay-censored data, for example to model median change in vRNA. With this approach, there are no assumptions concerning the distribution of the errors in the model. However, there is an assumption that the median change has linear associations with continuous covariates in the model, including baseline vRNA.

At Day 3, the adjusted difference between arms in median change from baseline was -1.17 log_10_ copies/ml (95% CI: -2.42, 0.07) favoring the active arm. This is reasonably similar to the adjusted difference in mean change of -0.97 log_10_ copies/ml obtained from tobit regression, though estimated without making the assumption of normally distributed errors. There is a somewhat narrower CI for the difference in means, versus difference in medians, reflecting the gain in precision from assuming a normal distribution for the errors. At Day 7, the adjusted difference in median change was -0.96 log_10_ copies/ml, also favoring the active arm. However, it wasn’t possible to obtain a CI from the numerical methods used to fit the model, due to the high proportion of participants with vRNA <LLoQ at Day 7. At Day 14, the higher proportion with vRNA <LLoQ meant the difference in median change between arms couldn’t be estimated.

### Analyzing Repeated vRNA Over Time

#### 5. Imputed values can affect estimates from MMRM due to correlation structure

Another strategy in several recent COVID-19 trials has been to use an MMRM with single-imputation for vRNA values <LLoQ [2–14]. These models estimate the difference in mean vRNA change in each arm at each timepoint, in a similar manner as linear regression models fit separately by timepoint. However, MMRMs incorporate a stronger assumption about the distribution of errors across timepoints, specifically that they follow a multivariate normal distribution with a specified correlation structure. Using this assumption, a global test evaluating the null hypothesis of no difference between arms in vRNA change at any timepoint can be undertaken. The stronger assumption may provide improved precision in estimating the differences in mean change at each timepoint by borrowing information between timepoints. However, this assumption may not be appropriate when using singly-imputed values for measurements <LLoQ as the correlation structure is affected by imputation. As an example, participants with vRNA <LLoQ at Days 7 and 14 will have identical imputed changes at both timepoints leading to higher correlations of errors in the model, than if the actual values <LLoQ were observed. To illustrate the impact of this, Table 3E shows results from MMRMs for changes from baseline to Days 3, 7, and 14. Compared with the estimates from models fitted separately at each timepoint (Table 3B), the borrowing of information through the correlation structure leads to smaller estimated differences in mean change between arms, particularly at Day 3 and to a lesser extent at Day 7 for both imputation approaches. This attenuation is driven by including Day 14, where ∼90% of participants had vRNA <LLoQ; removal of this timepoint from the MMRM reduces the magnitude of the attenuation (Table 3F). The estimates remain biased, however, for the same reasons as those obtained from separate regression models at each timepoint.

Extensions to MMRM that account for censored data exist (also known as linear mixed effects models for censored responses[LMEC]), but still require the multivariate normality assumption[23,24]. A caveat with these models is that they can be difficult to implement in standard statistical software, especially as the number of timepoints increases. Estimated differences between arms in mean change from baseline to Days 3 and 7 from LMEC are shown in Table 3G. The estimates are similar to those from the tobit regression models fitted separately at Days 3 and 7 (Table 3C). The stronger multivariate normal assumption leads to small gains in precision at Day 7 as seen by the narrower CI, though the gain at Day 3, where there’s less censoring, is negligible. As with the separate regression models, we didn’t pursue LMEC over the three days, as the high level of censoring at Day 14 meant that a normality assumption couldn’t be reasonably verified.

### Analyzing Proportion of Participants with vRNA <LLoQ Over Time

#### 6. Strategies that don’t rely on quantitative values may be preferred with large % <LLoQ

When there is a high proportion of participants with vRNA <LLoQ at one or more timepoints, it may be more appropriate to focus on how this proportion changes with time. This could be analyzed over time using log-binomial models fit using generalized estimating equations (GEE). However, due to problems with numerical algorithms, in ACTIV-2 we used Poisson regression models modified for binary outcomes [25] fit using GEEs with independence working correlation structure and robust standard errors, adjusting for baseline vRNA. When implementing this model across the three days, the proportion with vRNA <LLoQ didn’t differ between arms (Supplemental Table 2). When excluding the Day 14 measurements, where ∼90% of participants had vRNA <LLoQ, the results for Days 3 and 7 were almost identical, confirming this method isn’t sensitive to including timepoints with high proportions <LLoQ. This strategy can lead to loss in statistical power compared to analyses of quantitative vRNA, so is best reserved for when high proportions of participants are expected to have vRNA <LLoQ at one or more timepoints. However, there is also no need to restrict the analysis population to participants with vRNA ≥LLoQ, potentially providing more comprehensive analyses of qualitative vRNA in the overall study population.

## DISCUSSION

In this paper we summarize methods commonly used in outpatient COVID-19 therapeutic trials for analyzing quantitative changes in SARS-CoV-2 RNA over time, and through an illustrative example from the ACTIV-2 study, highlight potential pitfalls. In ACTIV-2, our primary virology analyses focused on comparing the proportion of participants with vRNA <LLoQ over time, and examined vRNA levels rather than changes. As the pandemic has evolved and we have learned more about viral trajectories and variability, so has our thinking about the best analytic strategy. Since designing ACTIV-2, we have implemented exploratory analyses examining treatment effects on changes in vRNA over time using tobit regression models with adjustment for baseline RNA, restricted to participants with baseline vRNA ≥LLoQ, a method we advocate for in this paper [19,26,27].

In our illustrative example, the primary focus was on the population with quantifiable vRNA at baseline, which has been a focus in recent COVID-19 studies. This was reasonable in our analysis as none who were <LLoQ at baseline had quantifiable vRNA at later timepoints. Including these individuals in analyses using imputed values would have led to imputed changes of zero and likely attenuation of the estimated mean changes. Regression analyses for censored data are more complex if such individuals are included, requiring strong, unverifiable assumptions about the distribution of vRNA changes over time among those with baseline vRNA <LLoQ. Looking more broadly across the study population in phase II placebo-controlled evaluations in ACTIV-2 (N=1565 enrolled with a median of 6 days from symptom onset), we observed that only 14% (of 287) of those with vRNA <LLoQ at baseline later had quantifiable vRNA. As new studies are developed, potentially with enrollment closer to onset of symptoms, the decision to exclude those <LLoQ at baseline should be carefully scrutinized, as doing so could remove individuals on an upward viral load trajectory and we lack understanding of these trajectories in the setting of vaccination, reinfection, and emergent variants. At a minimum, documenting viral shedding changes among participants with baseline vRNA <LLoQ is important, and analyses stratified by level (<LLoQ and ≥LLoQ at baseline) might be pursued.

The methods considered in this paper aren’t exhaustive of imputation or modeling strategies, but were chosen to align with methods from recent publications of COVID-19 trials. We focus on single-imputation, and don’t evaluate the performance of multiple-imputation strategies, which are more complicated and rely on distributional assumptions to support the imputation, but may reduce potential biases with imputation highlighted in this paper [28,29]. We also haven’t evaluated the statistical performance of these methods through formal simulation studies, which may add further insights to benefits or downsides of the analytic strategies, particularly when high proportions of participants have vRNA <LLoQ during follow-up, where verification of model assumptions becomes more difficult. We also haven’t considered potential biases due to missing data, for example, missingness arising due to hospitalization, if hospitalized participants have higher vRNA levels. In designing studies, the impact on power and precision in estimating treatment effects needs consideration [30]. Finally, analysis of vRNA changes among participants with baseline levels above a threshold (e.g., the LLoQ) leads to estimated mean changes within each arm that are affected by regression to the mean, though estimated differences in mean changes between randomized arms are not. Despite these limitations, our paper highlights key issues and considerations when analyzing SARS-CoV-2 RNA data from outpatient treatment trials. These methods aren’t only applicable in the COVID-19 setting, but should be considered when analyzing any biomarker that is measured with an assay with an LLoQ.

## Recommendations

The best practices in analyzing SARS-CoV-2 RNA from outpatient trials depend on the number of timepoints and proportion of results <LLoQ. Regardless of the planned analysis, some key details should be reported to facilitate interpretation.

1. Provide sufficient details of the RT-qPCR assay, including the LLoQ.
2. Explain who is included in the analysis, such as via a CONSORT-type diagram (see Supplemental Figure 1), including an accounting of missing data and the reasons for missing (e.g., death, hospitalization, loss to follow-up, sample not obtained, sample processing/shipping issue).
3. If restricting the analysis population to those with quantifiable baseline vRNA, describe outcomes among those with vRNA <LLoQ.
4. Although we don’t recommend the use of single-imputation, if used, the choice of imputed values should be provided, and implications for interpretation of results discussed.
5. Include descriptive summaries of vRNA by treatment arm and timepoint. We suggest including two figures (see Figure 1): distributions of quantitative levels (e.g., box and whisker plots) and distribution of vRNA categories (e.g., <LLoQ versus ≥LLoQ).

Analytic strategies to estimate differences between arms we recommend:

1. Methods that address censoring without imputation, such as tobit or median regression, or LMEC [23,24] be prioritized. But with increased censoring:
  a. Normality assumptions underlying regression analysis for censored data cannot be evaluated over the full range of the distribution, and dropping timepoints with high levels of censoring from analysis may be appropriate.
  b. Differences in medians (and their CIs) between arms might not be estimable from quantile regression.
2. Alternatively, consider non-parametric tests to analyze quantitative vRNA, such as the censored version of the Wilcoxon test (Gehan-Wilcoxon) which is implementable in standard software as a stratified test to account for baseline vRNA.
3. Comparing the proportion of participants with vRNA <LLoQ between arms over time may be preferred if there are high amounts of censoring.
4. With early, frequent measurements (e.g., daily), more complex extensions of LMEC that evaluate viral dynamics (e.g., estimating initial increases and subsequent vRNA decay) [20,31–34], or time-to-viral clearance via methods for time-to-event data [4–7,10,35,36] might be used.

## Data Availability

Data are available under restricted access due to ethical restrictions. Access can be requested by submitting a data request at https://submit.mis.s-3.net/ and will require the written agreement of the AIDS Clinical Trials Group (ACTG) and the manufacturer of the investigational product. Requests will be addressed as per ACTG standard operating procedures. Completion of an ACTG Data Use Agreement may be required.

## ACKNOWLEDGMENTS

We thank the study participants, site staff, site investigators, and the entire ACTIV-2/A5401 study team; the AIDS Clinical Trials Group, including Lara Hosey, Jhoanna Roa, and Nilam Patel; the ACTG Laboratory Center, including Grace Aldrovandi and William Murtaugh; Frontier Science, including Marlene Cooper, Howard Gutzman, and Kevin Knowles; the Harvard Center for Biostatistics in AIDS Research (CBAR) and ACTG Statistical and Data Analysis Center (SDAC) including Ronald Bosch and Linda Harrison; the National Institute of Allergy and Infectious Diseases (NIAID) / Division of AIDS (DAIDS); Bill Erhardt; the Foundation for the National Institutes of Health and the Accelerating COVID-19 Therapeutic Interventions and Vaccines (ACTIV) partnership, including Stacey Adams; and the PPD clinical research business of Thermo Fisher Scientific. We also thank AstraZeneca for partnership in the ACTIV-2 study and for donation of study drug for the cohort used as the illustrative example in this analysis.

