## Supplemental Material for "Statistical Challenges when Analyzing SARS-CoV-2 RNA Measurements Below the Assay Limit of Quantification in COVID-19 Clinical Trials"

Supplemental Table 1: Summary of Analysis Timepoints, Assay Limits, and Choice of Imputed Values for Quantitative SARS-CoV-2 RNA Below Assay Lower Limit of Quantification in Recent Outpatient COVID-19 Trials 13

### SUPPLEMENTAL METHODS

#### Model Implementation Details and SAS Programming Code

Below is a snapshot of the first 12 observations used in the illustrative example, with descriptive details on the variables used in analysis. The following SAS programming code was used to implement each statistical model using SAS version 9.4.

| Obs | PUBLICID | treatment | baserna | basernacat | visitday | rna | rnacat | changebase | lower | censor | rightchangebase |
|---|---|---|---|---|---|---|---|---|---|---|---|
| 1 | 50070162 | 1 | 6.50 | >= LLOQ | 3 | 2.68 | >= LLOQ | -3.82 | -3.82 | 0 | 103.82 |
| 2 | 50070162 | 1 | 6.50 | >= LLOQ | 7 | 2.00 | < LLOQ | -4.50 | . | 1 | 104.50 |
| 3 | 50070162 | 1 | 6.50 | >= LLOQ | 14 | 2.00 | < LLOQ | -4.50 | . | 1 | 104.50 |
| 4 | 50120111 | 0 | 2.00 | < LLOQ | 3 | 2.00 | < LLOQ | 0.00 | . | 1 | 100.00 |
| 5 | 50120111 | 0 | 2.00 | < LLOQ | 7 | 2.00 | < LLOQ | 0.00 | . | 1 | 100.00 |
| 6 | 50120111 | 0 | 2.00 | < LLOQ | 14 | 2.00 | < LLOQ | 0.00 | . | 1 | 100.00 |
| 7 | 50140111 | 1 | 6.67 | >= LLOQ | 3 | . | | . | . | . | . |
| 8 | 50140111 | 1 | 6.67 | >= LLOQ | 7 | 2.52 | >= LLOQ | -4.15 | -4.15 | 0 | 104.15 |
| 9 | 50140111 | 1 | 6.67 | >= LLOQ | 14 | 2.00 | < LLOQ | -4.67 | . | 1 | 104.67 |
| 10 | 50140112 | 0 | 8.00 | >= LLOQ | 3 | 6.65 | >= LLOQ | -1.35 | -1.35 | 0 | 101.35 |
| 11 | 50140112 | 0 | 8.00 | >= LLOQ | 7 | 5.34 | >= LLOQ | -2.66 | -2.66 | 0 | 102.66 |
| 12 | 50140112 | 0 | 8.00 | >= LLOQ | 14 | 2.00 | < LLOQ | -6.00 | . | 1 | 106.00 |

*Variable Details and Data Dictionary*

| | |
|---|---|
| PUBLICID | Blinded participant ID variable for public dissemination |
| TREATMENT | Treatment Indicator variable: 1 = tixagevimab/cilgavimab (active) and 0 = placebo |
| BASERNA | Baseline viral RNA ($log_{10}$ copies/ml) |
| BASERNACAT | Categorical Baseline viral RNA:<br>'>= LLOQ' for quantifiable values and '< LLOQ' for values below the assay lower limit of quantification |
| VISITDAY | Post-Baseline Visit on day X, where X = 3, 7 or 14 days |
| RNA | Viral RNA ($log_{10}$ copies/ml) on day X (defined by **visitday**).<br>Results < LLoQ are imputed as 2 $log_{10}$ copies/ml (which was the LLoQ) |
| RNACAT | Categorical viral RNA at on day X (defined by **visitday**):<br>'>= LLOQ' for quantifiable values and '< LLOQ' for values below the assay lower limit of quantification |
| CHANGEBASE | Change in viral RNA ($log_{10}$ copies/ml) from baseline to day X (defined by **visitday**):<br>Post-Baseline RNA on day X – Baseline RNA |
| LOWER | Analysis variable used in regression models for censored data. Lower = viral RNA value when the post-baseline value is quantifiable, and lower = "." (i.e., missing) when the post-baseline value is <LLoQ |
| CENSOR | Indicator variable whether the post-baseline value is left-censored (i.e., <LLoQ):<br>1 = censored, 0 = not censored |
| RIGHTCHANGEBASE | Analysis variable used in quantile regression (as software is designed for right-censored data).<br>**changebase** is transformed to a right-censored variable, calculated as: 100 – **changebase** |

*Modified Poisson Regression with robust standard error*

The proportion <LLoQ can be compared between arms adjusting for baseline viral RNA using a binary regression approach, such as modified Poisson regression with robust standard error [1] fit with generalized estimating equations (GEE) using an independence working correlation structure.  We implement this model using PROC GENMOD with a Poisson distribution and log link function. The REPEATED statement is included to indicate that the viral RNA values are repeated in the dataset within person with ID number **publicid**, and TYPE=IND to use an independence working correlation structure.

```
PROC GENMOD DATA=dataset;
    CLASS publicid treatment visitday;
    MODEL censor = visitday treatment*visit baserna / DIST=POISSON LINK=LOG;
    REPEATED SUBJECT=publicid / TYPE=IND;
RUN;
```

*Analysis of Covariance (ANCOVA) with imputed values*

ANCOVA models can be implemented in SAS via several procedures.  One such procedure is PROC GLM and can be implemented using the following code, including a where statement to restrict to those ≥LLoQ at baseline and to specify a post-baseline visit on Day X.

```
PROC GLM DATA=dataset;
    WHERE basernacat = “>= LLOQ” and visitday = <insert>;
    CLASS treatment;
    MODEL changebase = treatment baserna / SOLUTION;
RUN;
```

*Linear regression for censored data (tobit regression)*

Linear regression for censored data can be implemented in SAS using PROC LIFEREG, with restrictions in the where statement to subset to the population with baseline viral RNA ≥LLoQ and to a specific visit on Day X.  The MODEL statement should reflect the left-censored nature of the viral RNA data, such that for each observation, the interval in the model (lower, upper) is the range of possible values the observation could take.  For results ≥LLoQ, lower and upper values are identical and equal to the quantifiable viral RNA value.  For results <LLoQ, the lower is set to missing (i.e., “.”) and the upper is set to the change in viral RNA from baseline to the LLoQ of 2 log$_{10}$ copies/ml (i.e., 2 – baseline).

```
PROC LIFEREG DATA=dataset;
        WHERE basernacat = “>= LLOQ” and visitday = <insert>;
        CLASS treatment;
        MODEL (lower, changebase)  = treatment baserna / DISTRIBUTION=NORMAL;
RUN;
```

*Median regression for censored data*

Quantile regression for survival data can be implemented in SAS using PROC QUANTLIFE (note that PROC QUANTREG is not appropriate in our setting as it requires all observations to be observed, so does not allow for censored measurements). To specify median regression, the 50th quantile should be indicated in the options of the MODEL statement. The outcome in the MODEL statement is composed of two pieces: the response variable and a variable indicating which observations are "censored" (note: here censored values are flagged as **censor**=1). Because this procedure has been designed to handle right-censored data, the response variable must be right censored in the analysis dataset. Therefore, we created an analysis variable called **rightchangebase** that is calculated as '100 – **changebase**' to ensure all values are right censored. Results need to be transformed back to the original scale of measurement for dissemination purposes.

```
PROC QUANTLIFE DATA=dataset;
        WHERE basernacat = “>= LLOQ” and visitday = <insert>;
        CLASS treatment;
        MODEL rightchangebase*censor(1)  = treatment baserna / QUANTILES=(0.50);
RUN;
```

*MMRM with imputed values*

Mixed effects models for repeated measures can be implemented in SAS via PROC MIXED. Here, we use a REPEATED statement to indicate that we have repeated viral RNA values across study visits within SUBJECT labeled by **publicid**, and use an unstructured covariance structure (shown by TYPE=UN). The MODEL statement contains an interaction term between treatment and visit to calculate the treatment differences at each time point. Note: the WHERE statement only restricts the analysis to those ≥LLoQ at baseline as all post-baseline timepoints are included in the same model. If we wanted to restrict the post-baseline timepoints to include only Days 3 and 7 (as we did in our illustrative example) we would add an additional restriction here.

```
PROC MIXED DATA=dataset;
    WHERE basernacat = “>= LLOQ”;
    CLASS publicid treatment visitday;
    MODEL changebase = visitday treatment*visitday baserna / SOLUTION;
    REPEATED visit / SUB=publicid TYPE=UN;
RUN;
```

*MMRM for censored data (LMEC)*

The programming for the linear mixed effects model for censored data (LMEC) is more complex to implement as we wanted to create a repeated measures model with an unstructured covariance matrix. We opted to manually implement the likelihood function within PROC NLMIXED using the following programming code. Note: this code is specifically designed for two timepoints as it implements a bivariate normal distribution.  The input dataset needs to be transposed so that for each time point (i.e., Day 3 and Day 7) there is a change in viral RNA and censor indicator for each person, and a new variable, **changes**, which is set to 2 if both Day 3 and Day 7 viral RNA are not missing, set to 1 if one viral RNA values is missing, and is set to missing if both Day 3 and Day 7 viral RNA are missing.

| Obs | PUBLICID | treatment | baserna | basernacat | chg3 | rna3 | censor3 | chg7 | rna7 | censor7 | changes |
|---|---|---|---|---|---|---|---|---|---|---|---|
| 1 | 50070162 | 1 | 6.50 | >= LLOQ | -3.82 | 2.68 | 0 | -4.50 | 2.00 | 1 | 2 |
| 2 | 50120111 | 0 | 2.00 | < LLOQ | 0.00 | 2.00 | 1 | 0.00 | 2.00 | 1 | 2 |
| 3 | 50140111 | 1 | 6.67 | >= LLOQ | . | . | . | -4.15 | 2.52 | 0 | 1 |
| 4 | 50140112 | 0 | 8.00 | >= LLOQ | -1.35 | 6.65 | 0 | -2.66 | 5.34 | 0 | 2 |
| 25 | 50760132 | 1 | 5.15 | >= LLOQ | . | . | . | . | . | . | . |

```
PROC NLMIXED DATA=dataset_transpose;

    WHERE basernacat = “>= LLOQ”;

    PARMS alpha3=3  alpha7=3  trt3=1  trt7=1  bsl3=3  bsl7=3  rho=0  sd3=1  sd7=1;

    BOUNDS 1 > rho > -1, sd3  sd7 > 0;

    pi = CONSTANT('pi');

    mu3 = alpha3 + bsl3*baserna + trt3*treatment;

    mu7 = alpha7 + bsl7*baserna + trt7*treatment;

    z3 = (chg3-mu3)/sd3;

    z7 = (chg7-mu7)/sd7;

    rhosq=rho*rho;
```

```
IF ((chg3 NE .) AND (chg7 NE .)) THEN DO;

    IF (censor3=0 AND censor7=0) THEN

    L = -log(2*pi) - log(sd3) - log(sd7) - (0.5*log(1-rhosq)) - ( (z3**2 - (2*rho*z3*z7) + z7**2) / (2*(1-rhosq)) );

    ELSE IF (censor3=0 AND censor7=1) THEN

    L = -0.5*log(2*pi) - log(sd3) - (z3*z3/2) + log(PROBNORM((chg7 - (mu7 + (rho*sd7*z3))) / (sd7*sqrt(1-rhosq))));

    ELSE IF (censor3=1 AND censor7=0) THEN

    L = -0.5*log(2*pi) - log(sd7) - (z7*z7/2) + log(PROBNORM((chg3 - (mu3 + (rho*sd3*z7))) / (sd3*sqrt(1-rhosq))));

    ELSE IF (censor3=1 AND censor7=1) THEN

    L = PROBBNRM(z3, z7, rho);

END;

ELSE IF((chg3 NE .) AND (chg7 EQ .)) THEN DO;

    IF (censor3 = 0)            THEN L = -0.5*(log(2*pi)) - log(sd3) - (z3*z3/2);

    ELSE IF (censor3 = 1)       THEN L = log(PROBNORM(z3));

END;
```

```
        ELSE IF ((chg3 EQ .) AND (chg7 NE .)) THEN DO;
                IF (censor7 = 0)            THEN L = -0.5*(log(2*pi)) - log(sd7) - (z7*z7/2);
                ELSE IF (censor7 = 1)       THENL = log(PROBNORM(z7));
        END;
        ELSE IF ((chg3 EQ .) AND (chg7 EQ .)) THEN DO;
                L = .;
        END;
        MODEL changes ~ GENERAL(L);
RUN;
```

### SUPPLEMENTAL TABLES AND FIGURES

**Supplemental Table 1: Summary of Analysis Timepoints, Assay Limits, and Choice of Imputed Values for Quantitative SARS-CoV-2 RNA Below Assay Lower Limit of Quantification in Recent Outpatient COVID-19 Trials**

| Study | Analysis Timepoints (Days) | Assay Limits | Imputation Method |
|---|---|---|---|
| ACTIV-2 [2] | 0[a], 3, 7, 14[b] | LLoQ: 2 $log_{10}$ copies/ml<br>LOD: 1.4 $log_{10}$ copies/ml | BLoQ and detectable → midpoint of LLoQ and LOD<br>BLoQ and undetectable → midpoint of LOD and zero |
| REGEN-COV and REGEN-COV2 [3,4] | 1[a], 7, 15, 29<br>1[a], 3, 5, 7 | LLoQ: 2.85 $log_{10}$ copies/ml | BLoQ and detectable → midpoint of LLoQ and zero<br>BLoQ and undetectable → 0 $log_{10}$ copies/ml |
| BLAZE-1 and BLAZE-4 [5–7] | 1[a], 3, 5, 7, 11 | Ct limit: 45 | Negative → equal to the Ct limit (0 $log_{10}$ copies/ml) |
| Molnupiravir [8] | 1[a], 3, 5, 7, 14 | LLoQ 3 $log_{10}$ copies/ml | BLoQ → equal to the LLoQ |
| TACKLE [9] | 1[a], 3, 6, 15, 29 | LLoQ: 3.348 $log_{10}$ copies/ml | BLoQ → midpoint of LLoQ and zero |
| COMET-ICE [10] | 1[a], 8 | LLoQ: 3.35 $log_{10}$ copies/ml<br>LOD: 3.17 $log_{10}$ copies/ml | BLoQ and detectable → midpoint of LLoQ and LOD<br>BLoQ and undetectable → midpoint of LOD and zero |
| Peginterferon Lambda-1a [11] | 0[a], 7, 10, 14 | Ct limit: 40 (i.e., LOD: 1.6 $log_{10}$ copies/ml) | BLoD → Ct = 42 (i.e., 1 $log_{10}$ copies/ml) |
| CONV-ERT [12] | 0[a], 7, 28 | LOD: 3 $log_{10}$ copies/ml | BLoD → equal to the limit |
| TOGETHER [13] | 0[a], 3, 7 or 0[a],1-14 | LOD: 1 $log_{10}$ copies/ml | BLoD → 0 $log_{10}$ copies/ml |

| | | | |
|---|---|---|---|
| STOIC [14] | 0[a], 7, 14 | Ct limit: 40 | BLoD → equal to the Ct limit |
| Hydroxychloroquine [15] | 1[a], 3, 7 | LOD 3 $\log_{10}$ copies/ml | BLoD → equal to the LOD |
| EPIC-HR [16] | 1[a], 3, 5, 10, 14 | LOD: 2 $\log_{10}$ copies/ml | BLoD → midpoint of LOD and zero |
| MOVE-OUT [17] | 1[a], 3, 5, 10, 15, 29 | LLoQ: 2.70 $\log_{10}$ copies/ml | BLoQ → 1 copy lower than LLoQ |
| Remdesivir [18] | 0[a], 2, 3, 7 | LLoQ: 3.35 log10 copies/ml<br>LOD: 3.17 log10 copies/ml | BLoQ and detectable → midpoint of LLoQ and zero<br>BLoQ and undetectable → midpoint of LOD and zero |
| Ensitrelvir [19] | 1[a], 2, 4, 6, 9, 14, 21 | LLoQ: 2.08 log10 copies/ml | BLoQ → equal to the limit |

[a]Baseline Timepoint; [b]Days 21 and 28 dropped from later protocol versions. BLoQ = Below Limit of Quantification; LLoQ = Lower Limit of Quantification; LOD = Limit of Detection; BLOD = Below Limit of Detection; Ct = Cycle Threshold. Studies identified through PubMed search and review of COVID treatment guidelines.

**Supplemental Table 2: Proportion with RNA <LLoQ adjusting for Baseline RNA. Risk Ratios, 95% CIs and p-values among those with baseline viral RNA ≥LLoQ**

| Global P-value[a] | Day 3 | Day 7 | Day 14 |
|---|---|---|---|
| G: GEE Model (Modified Poisson Regression) across all three days | | | |
| 0.86 | 1.00 (0.47, 2.13)<br>p=0.99 | 1.20 (0.81, 1.77)<br>p=0.36 | 1.08 (0.87, 1.33)<br>p=0.50 |
| H: GEE Model (Modified Poisson Regression) across Days 3 and 7 | | | |
| 0.71 | 1.02 (0.51, 2.03)<br>p=0.96 | 1.20 (0.79 1.82)<br>p=0.40 | - - |

[a]Global p-value based on Wald's test. Risk ratios obtained using Modified Poisson Regression with robust standard error, fit with generalized estimating equations and independence working correlation. This model is not restricted to those ≥LLoQ at baseline.

GEE = Generalized Estimating Equation; N/A = Not Applicable.

**Supplemental Figure 1: Consort Flow for Participants Included in Analysis**

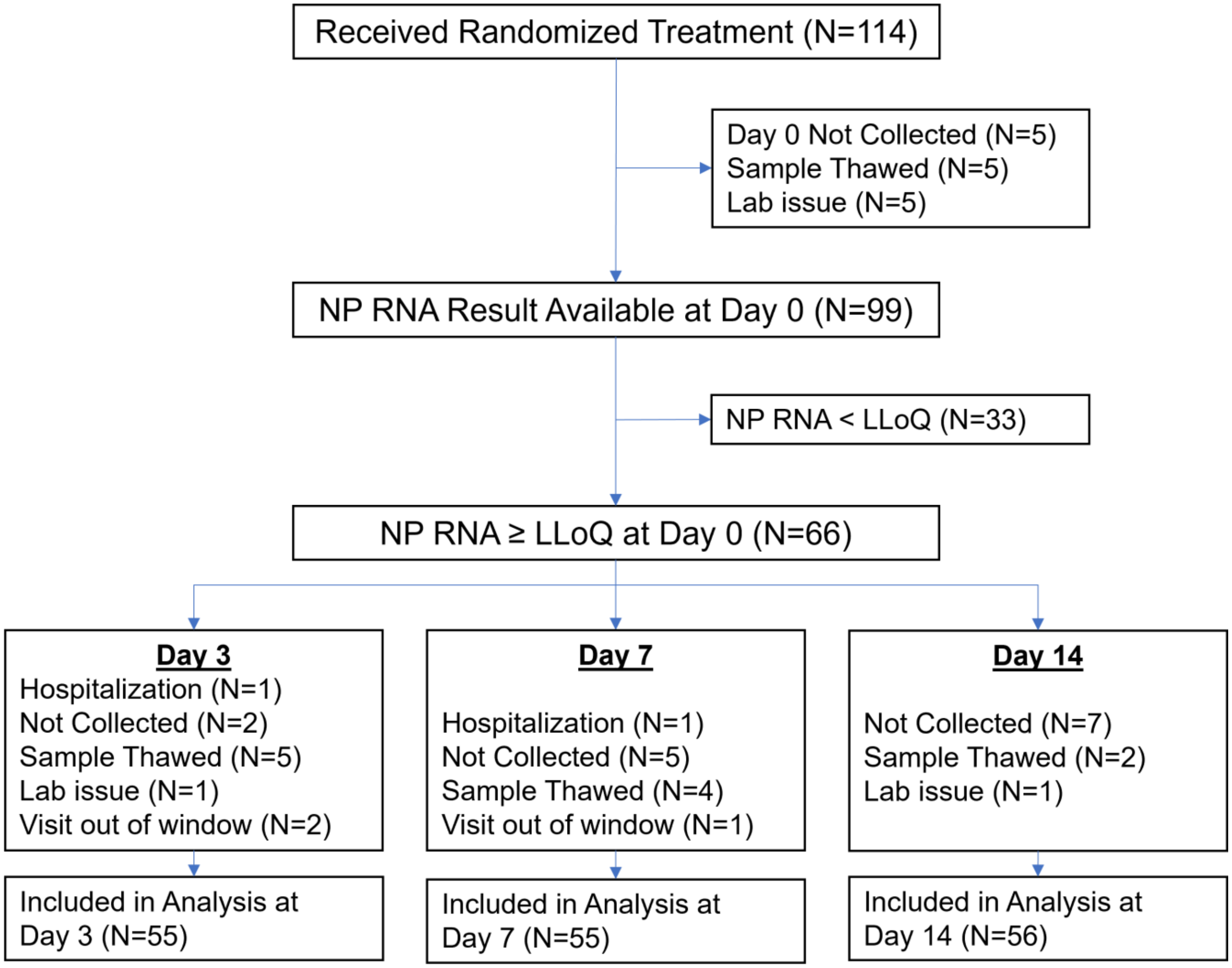

NP = Nasopharyngeal; LLoQ = Lower Limit of Quantification
